# Widespread Hyperalgesia Predicts Mortality in Pancreatic Adenocarcinoma

**DOI:** 10.64898/2026.05.19.26353594

**Authors:** Mahya Faghih, Marko Damm, Marie-Theres Kassik, Lara Cheesman, Sophie Rauschenberg, Søren S Olesen, Daniel A Laheru, Lei Zheng, Anna E Phillips, Dhiraj Yadav, Asbjørn M Drewes, Jonas Rosendahl, Vikesh K Singh, the International Pancreatic Pain Consortium

## Abstract

Pain in pancreatic ductal adenocarcinoma (PDAC) is associated with poor survival, but whether altered pain processing carries prognostic significance is unknown. We analyzed a prospective cohort of 143 patients with PDAC who underwent pancreatic quantitative sensory testing (P-QST) after diagnosis. Patients were classified as having normal pain processing (n=84), segmental hyperalgesia (n=30), or widespread hyperalgesia (n=29). Survival was measured from the date of P-QST assessment. During follow-up, 70 deaths occurred. Widespread hyperalgesia was associated with increased mortality in unadjusted Cox analysis (HR 1.96, 95% CI 1.14–3.35) and after adjustment for age, sex, tumor stage, comorbidity, opioid treatment, and body mass index (adjusted HR 2.33, 95% CI 1.30–4.15). Segmental hyperalgesia was not associated with mortality. Kaplan–Meier analysis demonstrated lower survival probability in the widespread hyperalgesia group (log-rank p=0.025). These findings suggest that widespread hyperalgesia, reflecting altered central pain processing, identifies a subgroup of PDAC patients at increased risk of mortality independent of conventional clinical factors.

## Introduction

Pancreatic ductal adenocarcinoma (PDAC) is frequently accompanied by abdominal pain, which contributes substantially to impaired quality of life and functional decline.[29] Pain is reported in approximately 60% of patients at or before diagnosis and increases up to 80% with disease progression; it is often refractory to standard analgesic strategies, including opioid-based regimens and neurolytic interventions.[18,29] Beyond symptom burden, presence and severity of abdominal pain in PDAC has been associated with adverse clinical outcomes, including reduced survival, although the mechanisms underlying this association remain incompletely understood.[14,19,22]

Cancer-related pain is increasingly recognized as a complex, multidimensional phenomenon that extends beyond local tumor effects.[9,20] In PDAC, pain has traditionally been attributed to peripheral mechanisms such as perineural invasion[13,32,33], neuroinflammation, pancreatic duct obstruction, and involvement of adjacent visceral and somatic structures. Neurotrophic signaling within the tumor microenvironment, including nerve growth factor (NGF), may contribute to neural remodeling and increased tumor innervation, further amplifying peripheral nociceptive input. [13,32,33] However, accumulating evidence from both malignant and benign pain conditions suggests that sustained nociceptive input can lead to altered central pain processing, resulting in amplified pain perception and generalized hypersensitivity.[1,7,9] Such alterations are commonly conceptualized as central sensitization and may have implications that extend beyond pain severity alone.

Quantitative sensory testing (QST) provides a standardized approach to characterize pain processing by probing peripheral and central nociceptive pathways. Pancreatic quantitative sensory testing (P-QST) was originally developed to study pain mechanisms in chronic pancreatitis. It leverages the viscerosomatic convergence of pancreatic and somatic afferents from relevant dermatomes together with assessment of pain modulation to characterize pancreas-related sensitization. Using a predefined algorithm, P-QST can classify patients into distinct sensory phenotypes, including normal pain processing, segmental hyperalgesia confined to pancreatic dermatomes, and widespread hyperalgesia reflecting more generalized sensitization.[8,23]

In prior work, we demonstrated that patients with painful, treatment-naïve PDAC exhibit abnormal pain processing compared with healthy controls, with increased prevalence of segmental and widespread hyperalgesia accompanied by lower pressure pain thresholds, enhanced temporal summation, and reduced pain tolerance.[7] Whether sensory phenotypes identified by P-QST in PDAC carry prognostic significance beyond pain characterization has not been examined.

Therefore, we aimed to investigate whether sensory phenotyping, as assessed by P-QST, is associated with all-cause mortality in patients with PDAC. We hypothesized that widespread hyperalgesia would be independently associated with higher mortality compared with normal pain processing.

## Methods

### Study design and participants

This study included patients with histologically confirmed PDAC who underwent pancreatic P-QST as part of a prospective observational cohort. Patients were enrolled from two tertiary pancreatic referral centers: The Johns Hopkins Medical Institutions (Baltimore, MD, USA) and University Hospital Halle (Halle, Germany). Eligible participants were ≥18 years of age and included patients with and without abdominal pain. Pancreatic-type pain was defined as pain located in the epigastric or upper abdominal region, with or without radiation to the back, and could be constant or intermittent.

Patients were excluded if they had undergone prior pancreatic resection, radiotherapy, or more than three chemotherapy cycles, to reduce potential confounding by treatment-related neuropathy[3,11], or if they had medical or neurological conditions that could interfere with quantitative sensory testing, including known peripheral neuropathy or other disorders affecting somatosensory function. Additional exclusion criteria included recent abdominal surgery and conditions judged by the investigators to compromise the validity or safety of sensory testing. All participants provided written informed consent prior to study participation. Institutional Review Board (IRB) approval was obtained at each study site (Johns Hopkins IRB 00143375 and Martin Luther University Halle/Wittenberg IRB 2021–189). Patients and/or the public were not involved in the design, conduct, reporting, or dissemination plans of this research.

### Clinical assessment and covariates

Baseline demographic and clinical data were collected at the time of P-QST assessment. Recorded variables included age, sex, body mass index (BMI), comorbidity burden assessed using the Charlson comorbidity index(CCI), tumor stage according to the Union for International Cancer Control (UICC) classification, presence of pancreatic-type pain and current analgesic treatment, including opioid use.

### Pancreatic quantitative sensory testing (P-QST)

P-QST was performed by trained study personnel using standardized equipment and a predefined testing sequence, as previously described. The protocol was designed to assess peripheral and central pain processing by exploiting viscerosomatic convergence between pancreatic afferents and somatic dermatomes together with general measures of neuronal excitability and descending (brainstem-spinal) modulation of pain.[23]

### Static quantitative sensory testing

Static testing included assessment of pressure pain detection thresholds using a handheld pressure algometer with a 1.0 cm² probe. Measurements were obtained on the participant’s dominant side at five standardized dermatomal sites: C5 (clavicular region), T10 dorsal (pancreatic viscerotome), T10 ventral (upper abdominal region), L1 (anterior superior iliac crest), and L4 (quadriceps region). Pressure thresholds were recorded in kilopascals. Summary measures were derived from pressure thresholds across tested sites, and ratios comparing pancreatic to non-pancreatic dermatomes were calculated according to the established protocol.[23]

### Dynamic quantitative sensory testing

Dynamic testing evaluated temporal summation and conditioned pain modulation (CPM). Temporal summation, a psychophysical measure of activity-dependent facilitation in nociceptive pathways reflecting increased spinal neuronal excitability (central sensitization), was assessed using repetitive pinprick stimulation. Identical pinprick stimuli were applied sequentially at fixed intervals to the T10 ventral dermatome and to a control site on the forearm. Participants rated pain intensity after a single stimulus and again after a series of repeated stimuli. Temporal summation was defined as the increase in perceived pain intensity during the repeated stimulus series compared with the single stimulus, reflecting progressive amplification of nociceptive signaling in the spinal cord.[23]

CPM as an indirect measure of descending pain modulation was assessed using a cold pressor paradigm, in which participants immersed their dominant hand in ice-cold water for a maximum predefined duration. Pressure pain tolerance was measured at L4 on non-dominant thigh before and immediately after the conditioning stimulus, and CPM capacity was calculated as the relative change in pressure pain tolerance following cold exposure. Cold pressor endurance time was recorded as an additional measure of pain tolerance.[23]

### Sensory phenotype classification

Based on predefined criteria and an established algorithm[8,23], participants were classified into one of three P-QST phenotypes: normal pain processing, segmental hyperalgesia confined to pancreatic dermatomes, or widespread hyperalgesia extending beyond the pancreatic region. This classification approach has been previously validated in chronic pancreatitis and applied consistently across studies.[8,23] No sensory testing results were used to guide clinical management.

### Mortality follow-up

All-cause mortality during follow-up was ascertained using institutional clinical records and follow-up data available within the participating institutions. Because P-QST defines baseline exposure status, survival time was measured from the date of P-QST assessment. The date of last follow-up was 09-21-2025.

### Statistical analysis

Data are presented as mean (SD), median (IQR), or n (%). Group comparisons used ANOVA or Kruskal–Wallis tests for continuous variables and χ² or Fisher’s exact tests for categorical variables, as appropriate. Overall survival was analyzed using Kaplan–Meier curves, the log-rank test, and Cox regression. Multivariable Cox regression models were adjusted for age, sex, tumor stage, comorbidity, opioid treatment, and body mass index. A p-value <0.05 was considered statistically significant. All statistical analyses were performed using Stata 18 (StataCorp, College Station, TX).

## Results

### Study cohort and P-QST phenotype distribution

A total of 143 patients with PDAC underwent P-QST between 2022-2025 and were classified into three predefined sensory phenotypes: normal pain processing (n = 84;59%), segmental hyperalgesia (n = 30; 31%), and widespread hyperalgesia (n = 29; 20%). P-QST was performed a median of 22 days (IQR 9–48) after PDAC diagnosis. The cohort was predominantly male (78/143; 55%) with a median age of 67 years (IQR 59–75) and a mean BMI of 25.1 kg/m² (SD 4.8). Advanced disease was common: 87 patients (61%) presented with UICC stage IV and an additional 13 (9%) with stage III, while 30 (21%) had stage II and 13 (9%) had stage I disease. Pancreatic-type pain was reported by 122 patients (85%) overall, and 51 patients (36%) were receiving opioid analgesics at the time of P-QST.

Baseline demographic and clinical characteristics, stratified by P-QST phenotype, are summarized in Table 1. No significant differences were observed across phenotypes for sex (p=0.431), age (p=0.716), BMI (p=0.513), CCI category (p=0.789), UICC tumor stage (p=0.328), prevalence of pancreatic-type pain (p=0.806), or baseline opioid use (p=0.263). Although opioid use was numerically more frequent in the widespread hyperalgesia group (14/29; 48%) than in the segmental (9/30; 30%) and normal (28/84; 33%) groups, this difference did not reach statistical significance.

**Table 1.** Demographic and clinical characteristics in the total patient cohort and stratified by P-QST phenotype.

|  | Total<br>(n=143) | Normal<br>(n=84) | Segmental<br>(n=30) | Widespread<br>(n=29) | P-value |
| --- | --- | --- | --- | --- | --- |
| Male sex, n (%) | 78 (55) | 43 (51) | 16 (53) | 19 (66) | 0.431 |
| Age, years (IQR) | 67 (59-75) | 68 (59-77) | 63 (57-72) | 68 (61-73) | 0.716 |
| BMI, kg/m<sup>2</sup> (SD) | 25.1 (4.8) | 25.0 (4.9) | 24.6 (4.2) | 26.0 (5.1) | 0.513 |
| Pancreatic pain, n (%) | 122 (85) | 70 (83) | 26 (87) | 26 (90) | 0.806 |
| Opioid treatment, n (%) | 51 (36) | 28 (33) | 9 (30) | 14 (48) | 0.263 |
| CCI category, n (%) |  |  |  |  |  |
| 0 | 47 (33) | 29 (35) | 8 (27) | 10 (34) | 0.789 |
| 1-2 | 65 (45) | 38 (45) | 13 (43) | 14 (48) |  |
| >2 | 31 (22) | 17 (20) | 9 (30) | 5 (17) |  |
| Tumor stage (UICC) |  |  |  |  |  |
| I | 13 (9) | 6 (7) | 6 (20) | 1 (3) | 0.328 |
| II | 30 (21) | 18 (21) | 7 (23) | 5 (17) |  |
| III | 13 (9) | 8 (10) | 1 (3) | 4 (14) |  |
| IV | 87 (61) | 52 (62) | 16 (53) | 19 (66) |  |

BMI, body mass index; CCI, Charlson comorbidity index; UICC, Union for International Cancer Control

### All-cause mortality by P-QST phenotype

Over a median follow-up of 273 days (IQR 121–365), 70 deaths occurred in the cohort. Kaplan–Meier survival curves demonstrated a significant separation between P-QST phenotypes, with the lowest survival probability observed in patients with widespread hyperalgesia (log-rank p = 0.025; Figure 1). Patients with segmental hyperalgesia exhibited survival trajectories comparable to those with normal pain processing throughout follow-up. Numbers at risk over time are shown below the survival curves and indicate a more rapid decline in the widespread hyperalgesia group compared with the other phenotypes.

**Figure 1.**
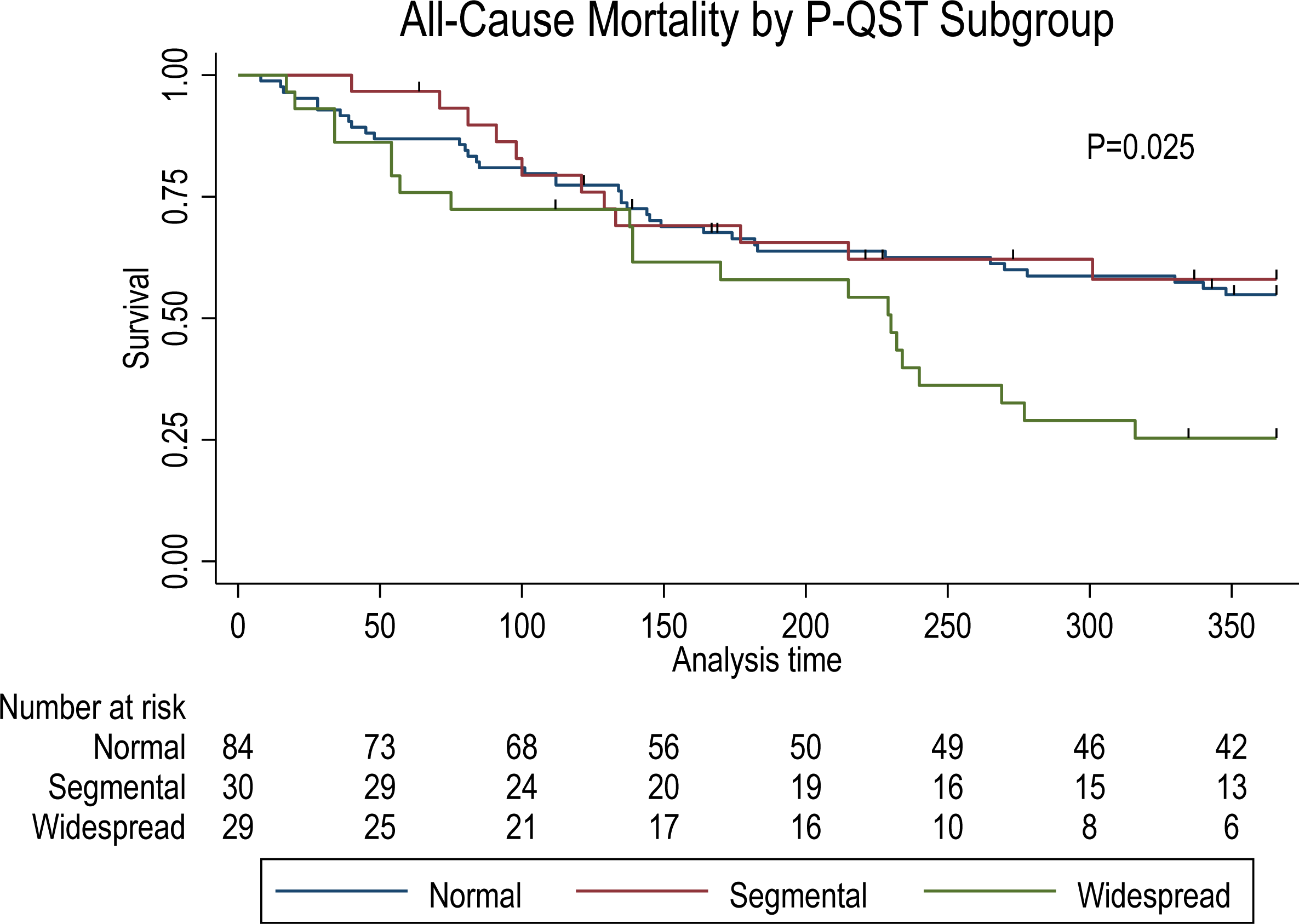
All-cause mortality by P-QST phenotypes in pancreatic ductal adenocarcinoma.

In unadjusted Cox regression, widespread hyperalgesia was associated with a nearly two-fold increase in all-cause mortality compared with normal pain processing (HR 1.96, 95% CI 1.14–3.35; p=0.014). The unadjusted HR for segmental hyperalgesia versus normal pain processing did not reach statistical significance.

After adjustment for age, sex, BMI, UICC tumor stage, CCI category, and baseline opioid use, the association between widespread hyperalgesia and mortality strengthened modestly (adjusted HR 2.33, 95% CI 1.30–4.15; p=0.004; Table 2). The adjusted HR for segmental hyperalgesia versus normal pain processing remained non-significant.

**Table 2.** All-cause mortality during follow-up stratified by P-QST phenotype.

|  | n | Follow-up<br>(person-years) | Deaths<br>(n) | HR – unadjusted<br>(95% CI) | P-value | HR – adjusted*<br>(95% CI) |
| --- | --- | --- | --- | --- | --- | --- |
| Normal | 84 | 57.6 | 37 | 1.00 (reference) | - | 1.00 (reference) |
| Segmental | 30 | 20.3 | 12 | 0.92 (0.48-1.76) | 0.925 | 1.01 (0.52-1.97) |
| Widespread | 29 | 15.9 | 21 | 1.96 (1.14-3.35) | 0.014 | 2.33 (1.30-4.15) |

\*The model was adjusted for age, sex, tumor stage, comorbidity, opioid treatment and BMI

## Discussion

In this cohort of 143 patients with PDAC who underwent P-QST after confirmed diagnosis and before or early in the course of treatment, we found that patients with widespread hyperalgesia had significantly higher all-cause mortality compared with patients having normal pain processing. This association was present in unadjusted analyses and remained robust after adjustment for age, sex, tumor stage, comorbidity burden, opioid treatment, and BMI. In contrast, segmental hyperalgesia was not associated with mortality in either unadjusted or adjusted models. Taken together, these findings suggest that widespread hyperalgesia, reflecting a generalized sensitized pain processing phenotype, identifies a subgroup of PDAC patients with adverse prognosis.

Pain is common in PDAC[18,29] and has long been associated with reduced survival[14,19,22], yet prognostic assessment has largely relied on symptom reports rather than objective characterization of pain mechanisms. Symptom-based measures remain clinically meaningful but are inherently heterogeneous, reflecting tumor burden, psychosocial factors, treatment exposure, and prescribing practices. Despite this heterogeneity, pain and symptom burden in PDAC have shown independent prognostic significance even after adjusting for established disease markers. In the landmark study by Kelsen and colleagues, the presence of pain was an independent predictor of poorer survival in operable pancreatic cancer, including among patients who underwent resection.[14] In metastatic PDAC, pain has also remained an independent prognostic factor in multivariable models alongside established markers of advanced disease.[19] By contrast, P-QST phenotyping aims to characterize pain processing through standardized assessments of somatosensory function leveraging viscerosomatic convergence. The present results suggest that pain processing phenotypes, particularly widespread hyperalgesia, may provide prognostic information beyond clinical staging and conventional baseline features.

A notable aspect of our findings is the specificity of the association to widespread hyperalgesia. Although simplified, segmental hyperalgesia is generally thought to reflect sensitization largely confined to pancreas-related dermatomes and may be more directly attributable to localized nociceptive input.[8,17,23] In contrast, widespread hyperalgesia indicates hypersensitivity, central reorganization, and more generalized nociplastic pain processing beyond the segmental distribution. [8,17,23] The observation that only the widespread phenotype predicted mortality is; therefore, consistent with the possibility that generalized sensitization captures broader dimensions of disease impact or host response that are not fully reflected by tumor stage alone. Importantly, the lack of significant differences in measured baseline characteristics across phenotypes, together with persistence of the association after multivariable adjustment, argues against simple confounding by the included covariates.

Biological plausibility for a link between altered pain processing and PDAC outcomes is increasingly supported by mechanistic and translational evidence.[30,32,33] Peripheral mechanisms of PDAC-related pain, including perineural invasion, neuroinflammation, pancreatic duct obstruction, and tumor infiltration of adjacent structures are well established, and additional mechanisms such as activation of transient receptor potential (TRP)channels and perineural mast cell infiltration have been implicated in nociceptive signaling.[5,27,33] Beyond the periphery, central neuroinflammatory pathways have been described in PDAC-related pain, including microglial activation within central visceral sensory circuits.[4] Emerging tumor–nerve–immune frameworks further suggest that nociceptor signaling may shape the tumor microenvironment. Interactions between nociceptor neurons and cancer-associated fibroblasts, involving neuropeptides and growth factors such as calcitonin gene-related peptide (CGRP) and NGF, have been implicated in suppression of natural killer (NK) cell infiltration and function, with potential relevance to both tumor progression and pain intensity.[30] Complementary stromal–neural interactions, including myofibroblast-induced neuroplasticity and sympathetic nerve–fibroblast crosstalk, may further contribute to neural remodeling in pancreatic cancer.[21,25] Beyond these indirect pathways, pancreatic cancer cells can directly interact with sensory neurons through synapse-like structures, enabling glutamate-mediated signaling that promotes tumor growth and neural remodeling.[24] Notably, nociceptive innervation has been reported to correlate positively with pain intensity and inversely with NK-cell infiltration[30] and has been reported as an independent prognostic factor for overall and relapse-free survival[26]. While our study does not test these pathways directly, these lines of evidence support the concept that nervous system alterations captured by widespread hyperalgesia may reflect biologically meaningful processes relevant to outcome.

Clinically, the findings suggest that P-QST phenotyping could support early identification of a subgroup with high-risk pain processing soon after diagnosis and prior to substantial treatment exposure. If validated in an independent cohort, widespread hyperalgesia could be considered for use as a stratification factor in future observational studies and interventional trials, including studies evaluating mechanistically informed analgesic strategies and structured supportive care pathways. At the same time, it is essential to emphasize that our data demonstrates prognostic association rather than causality. Whether modifying pain processing or improving pain control can influence survival cannot be inferred from this study and requires prospective investigations.

The relationship between pain management and survival in PDAC is complex. Reported associations between opioid exposure and survival are inconsistent and likely confounded by indication-patients healthy enough to receive opioids may have better performance status and access to cancer-directed therapies.[16,28,35] In our cohort, opioid use was more frequent among patients with widespread hyperalgesia; however, widespread hyperalgesia remained associated with mortality after adjustment for opioid use, suggesting that the prognostic signal is not explained by baseline opioid use alone. However, opioid exposure was captured only as a baseline treatment variable without information on dose, duration, or escalation. We, therefore, could not evaluate dose–response relationships or opioid-induced hyperalgesia as contributors to the widespread phenotype or mortality. Similar concerns apply to celiac plexus neurolysis and related neurolytic procedures: although these interventions can improve pain control and reduce opioid use, their reported effects on survival have been mixed across randomized and observational studies.[10,31,34] By contrast, early palliative care has shown more consistent benefit for symptom burden and quality of life [6,12,15] and, in some studies, survival [2,6]. Accordingly, our findings should not be interpreted as supporting any specific analgesic intervention; rather, they suggest that widespread hyperalgesia may identify a subgroup with greater clinical and prognostic vulnerability who may benefit from earlier multidisciplinary supportive care within an evidence-based pain management approach.

Several limitations should be acknowledged. First, although we adjusted for major clinical covariates, residual confounding remains possible. Although additional data were available (including post–P-QST cancer-directed treatments and baseline clinical status measures such as performance status), these were not included in the current models; these factors could influence survival and could plausibly differ by phenotype. Second, P-QST phenotyping was performed at a single time point; whether widespread hyperalgesia is stable, evolves with disease progression, or is modified by oncologic treatment (including potential reductions in nociceptive input with tumor response) and analgesic escalation in PDAC remains unknown. Third, opioid use was included as a covariate, but opioid exposure is an imperfect proxy for analgesic burden and may reflect unmeasured pain severity or clinical decision-making; the direction and magnitude of any opioid-related confounding cannot be fully resolved here. Fourth, the widespread hyperalgesia subgroup was modest in size, and external validation will be critical prior to clinical translation. Fifth, patients well enough to undergo P-QST may not be fully representative of the broader PDAC population, particularly those with the poorest performance status, introducing potential selection bias relevant to mortality outcomes. Finally, we evaluated all-cause rather than cause-specific mortality, and the contribution of competing causes of death cannot be determined from these data.

In conclusion, we demonstrate that widespread hyperalgesia assessed by P-QST was independently associated with increased all-cause mortality in PDAC, beyond tumor stage and other measured clinical factors. These results support the concept that pain phenotypes capture clinically relevant information beyond symptom reporting and provide a rationale for future studies to validate the association, integrate treatment exposures and performance status into prognostic models, and investigate links between widespread hyperalgesia and tumor–nerve–immune biology suggested by emerging mechanistic data.

## Data Availability

The data underlying this study are not publicly available due to institutional and patient privacy restrictions but may be made available from the corresponding author upon reasonable request and appropriate ethical approval.

## Acknowledgments

## Author Contributions

Conceptualization M.F., M.D., A.M.D., S.S.O., V.K.S.; methodology M.F., M.D., J.R.; formal analysis S.S.O.; data curation M.F., M.D., L.C., S.R., M.T.K.; writing - original draft preparation M.F., M.D.,L.C.; writing – review and editing D.A.L., L.Z., A.E.P., A.M.D., S.S.O., V.K.S., J.R.; supervision A.M.D., S.S.O., V.K.S., All authors have read and agreed to the published version of the manuscript.

## References

[1] Bouwense SAW, Olesen SS, Drewes AM, Frokjaer JB, van Goor H, Wilder-Smith OHG. Is altered central pain processing related to disease stage in chronic pancreatitis patients with pain? An exploratory study. PLoS One 2013;8:e55460.

[2] Brugel M, Dupont M, Carlier C, Botsen D, Essi DE, Sanchez V, Slimano F, Perrier M, Bouche O. Association of palliative care management and survival after chemotherapy discontinuation in patients with advanced pancreatic adenocarcinoma: A retrospective single-centre observational study. Pancreatology 2023;23:403–410.

[3] Catalano M, Ramello M, Conca R, Aprile G, Petrioli R, Roviello G. Risk Factors for Nab-Paclitaxel and Gemcitabine-Induced Peripheral Neuropathy in Patients with Pancreatic Cancer. Oncology 2022;100:384–391.

[4] Chen K, Ye Q, Zhang Y, Qi Z, Huang Y, Lu W, Wang X, Wang Y, Cao L, Qiu S, Xu Y, Huang J, Xie J. CXCL1-CXCR2 signaling mediates the activation of microglia in the nucleus tractus solitarii to promote pancreatic cancer-induced pain. Brain Behav Immun 2025;123:1026–1041.

[5] Demir IE, Schorn S, Schremmer-Danninger E, Wang K, Kehl T, Giese NA, Algul H, Friess H, Ceyhan GO. Perineural mast cells are specifically enriched in pancreatic neuritis and neuropathic pain in pancreatic cancer and chronic pancreatitis. PLoS One 2013;8:e60529.

[6] Dufva I, Juhl G, Andersen F, Beier-Holgersen R, Maddocks M. Early Specialized Palliative Care for Unresectable Pancreatic Cancer: A Quasi-Experimental Study. J Pain Symptom Manage 2026;71:419–428.

[7] Faghih M, Damm M, Bandhauer C, Jager J, Olesen SS, Laheru DA, Zheng L, Phillips AE, Yadav D, Drewes AM, Rosendahl J, Singh VK. Pancreatic adenocarcinoma patients with pain have abnormal central pain processing. Pancreatology 2025;25:426–432.

[8] Faghih M, Phillips AE, Kuhlmann L, Afghani E, Drewes AM, Yadav D, Singh VK, Olesen SS, Pancreatic Quantitative Sensory Testing (P-QST) Consortium. Pancreatic QST Differentiates Chronic Pancreatitis Patients into Distinct Pain Phenotypes Independent of Psychiatric Comorbidities. Clin Gastroenterol Hepatol 2022;20:153–161.e2.

[9] Fitzcharles M, Cohen SP, Clauw DJ, Littlejohn G, Usui C, Hauser W. Nociplastic pain: towards an understanding of prevalent pain conditions. Lancet 2021;397:2098–2110.

[10] Fujii-Lau LL, Bamlet WR, Eldrige JS, Chari ST, Gleeson FC, Abu Dayyeh BK, Clain JE, Pearson RK, Petersen BT, Rajan E, Topazian MD, Vege SS, Wang KK, Wiersema MJ, Levy MJ. Impact of celiac neurolysis on survival in patients with pancreatic cancer. Gastrointest Endosc 2015;82:46–56.e2.

[11] Goldstein D, Von Hoff DD, Moore M, Greeno E, Tortora G, Ramanathan RK, Macarulla T, Liu H, Pilot R, Ferrara S, Lu B. Development of peripheral neuropathy and its association with survival during treatment with nab-paclitaxel plus gemcitabine for patients with metastatic adenocarcinoma of the pancreas: A subset analysis from a randomised phase III trial (MPACT). Eur J Cancer;52:85–91.

[12] Haun MW, Estel S, Rucker G, Friederich H, Villalobos M, Thomas M, Hartmann M. Early palliative care for adults with advanced cancer. Cochrane Database Syst Rev 2017;6:CD011129.

[13] Jiang L, Cai S, Weng Z, Zhang S, Jiang S. Peripheral, central, and chemotherapy-induced neuropathic changes in pancreatic cancer. Trends Neurosci 2025;48:124–139.

[14] Kelsen DP, Portenoy R, Thaler H, Tao Y, Brennan M. Pain as a predictor of outcome in patients with operable pancreatic carcinoma. Surgery 1997;122:53–59.

[15] Kim CA, Lelond S, Daeninck PJ, Rabbani R, Lix L, McClement S, Chochinov HM, Goldenberg BA. The impact of early palliative care on the quality of life of patients with advanced pancreatic cancer: The IMPERATIVE case-crossover study. Support Care Cancer 2023;31:250–3.

[16] Koulouris A, Baio G, Clark A, Alexandre L. Opioid burden in patients with inoperable pancreatic adenocarcinoma and the development of a multivariable risk prediction model for opioid use: A retrospective cohort study. Pancreatology 2023;23:818–828.

[17] Kuhlmann L, Olesen SS, Gronlund D, Olesen AE, Phillips AE, Faghih M, Drewes AM. Patient and Disease Characteristics Associate With Sensory Testing Results in Chronic Pancreatitis. Clin J Pain 2019;35:786 – 793.

[18] McNearney TA, Digbeu BDE, Baillargeon JG, Ladnier D, Rahib L, Matrisian LM. Pre-Diagnosis Pain in Patients With Pancreatic Cancer Signals the Need for Aggressive Symptom Management. Oncologist 2023;28:e1185–e1197.

[19] Morizane C, Okusaka T, Morita S, Tanaka K, Ueno H, Kondo S, Ikeda M, Nakachi K, Mitsunaga S. Construction and validation of a prognostic index for patients with metastatic pancreatic adenocarcinoma. Pancreas 2011;40:415–421.

[20] National Comprehensive Cancer Network. NCCN Clinical Practice Guidelines in Oncology (NCCN Guidelines®):Pancreatic Adenocarcinoma. 2025.

[21] Nigri J, Lan W, Fung ML, Kayser C, Deschenes A, Hinds J, Kaushalya S, Pawlak SA, Thalappillil JS, Nadella S, Hilmi M, Park W, Kappagantula R, Park Y, Zhao Z, Preall J, Iacobuzio-Donahue CA, Tracey KJ, Borniger JC, Tuveson DA. Myofibroblasts induce neuroplasticity to promote pancreatic inflammation and cancer progression. Cancer Discov 2026.

[22] Okusaka T, Okada S, Ueno H, Ikeda M, Shimada K, Yamamoto J, Kosuge T, Yamasaki S, Fukushima N, Sakamoto M. Abdominal pain in patients with resectable pancreatic cancer with reference to clinicopathologic findings. Pancreas 2001;22:279–284.

[23] Phillips AE, Faghih M, Kuhlmann L, Larsen IM, Drewes AM, Singh VK, Yadav D, Olesen SS, Pancreatic Quantitative Sensory Testing (P-QST) Consortium. A clinically feasible method for the assessment and characterization of pain in patients with chronic pancreatitis. Pancreatology 2020;20:25–34.

[24] Ren L, Liu C, Cifcibasi K, Ballmann M, Rammes G, Mota Reyes C, Tokalov S, Klingl A, Grunert J, Goyal K, Neckel PH, Mattheus U, Schoeps B, Yildizhan SE, Sezerman OU, Cevik NC, Sever EA, Karakas D, Safak O, Steiger K, Muckenhuber A, Gorgulu K, Chen Z, Zhang J, Ye L, Maula Ali MI, Tiwari VK, Romanyuk N, Giesert F, Saur D, Rad R, Schmid RM, Algul H, Kruger A, Friess H, Ceyhan GO, Istvanffy R, Demir IE. Sensory neurons drive pancreatic cancer progression through glutamatergic neuron-cancer pseudo-synapses. Cancer Cell ;43:2241–2258.e8.

[25] Sattler AL, Diba P, Hawthorne K, Pelz C, Grieco J, Korzun T, Chong B, Kuykendall MJ, Sears RC, Marks DL, Sherman MH, Zimmers TA, Eksi SE. Sympathetic nerve-fibroblast crosstalk drives nerve injury, fibroblast activation, and matrix remodeling in pancreatic cancer. JCI Insight 2026.

[26] Schorn S, Demir IE, Haller B, Scheufele F, Reyes CM, Tieftrunk E, Sargut M, Goess R, Friess H, Ceyhan GO. The influence of neural invasion on survival and tumor recurrence in pancreatic ductal adenocarcinoma - A systematic review and meta-analysis. Surg Oncol 2017;26:105–115.

[27] Schwartz ES, Christianson JA, Chen X, La J, Davis BM, Albers KM, Gebhart GF. Synergistic role of TRPV1 and TRPA1 in pancreatic pain and inflammation. Gastroenterology 2011;140:1283–2.

[28] Steele GL, Dudek AZ, Gilmore GE, Richter SA, Olson DA, Eklund JP, Zylla DM. Impact of Pain, Opioids, and the Mu-opioid Receptor on Progression and Survival in Patients With Newly Diagnosed Stage IV Pancreatic Cancer. Am J Clin Oncol 2020;43:591–597.

[29] Stoop TF, Javed AA, Oba A, Koerkamp BG, Seufferlein T, Wilmink JW, Besselink MG. Pancreatic cancer. Lancet 2025;405:1182–1202.

[30] Wang K, Ni B, Xie Y, Li Z, Yuan L, Meng C, Zhao T, Gao S, Huang C, Wang H, Ma Y, Zhou T, Feng Y, Chang A, Yang C, Yu J, Yu W, Zang F, Zhang Y, Ji R, Wang X, Hao J. Nociceptor neurons promote PDAC progression and cancer pain by interaction with cancer-associated fibroblasts and suppression of natural killer cells. Cell Res 2025;35:362–380.

[31] Wong GY, Schroeder DR, Carns PE, Wilson JL, Martin DP, Kinney MO, Mantilla CB, Warner DO. Effect of neurolytic celiac plexus block on pain relief, quality of life, and survival in patients with unresectable pancreatic cancer: a randomized controlled trial. JAMA 2004;291:1092–1099.

[32] Xu J, Yao H, Wang J, Jin Y, Chang W, Li L, Zou L. Perineural invasion and the “cold” tumor microenvironment in pancreatic cancer: mechanisms of crosstalk and therapeutic opportunities. Front Immunol 2025;16:1650117.

[33] Xu W, Liu J, Zhang J, Lu J, Guo J. Tumor microenvironment crosstalk between tumors and the nervous system in pancreatic cancer: Molecular mechanisms and clinical perspectives. Biochim Biophys Acta Rev Cancer 2024;1879:189032.

[34] Zylberberg HM, Nagula S, Rustgi SD, Aronson A, Kessel E, Kumta NA, DiMaio CJ, Lucas AL. Celiac Plexus Neurolysis Is Associated With Decreased Survival in Patients With Pancreatic Cancer: A Propensity Score Analysis. Pancreas 2022;51:153–158.

[35] Zylberberg HM, Woodrell C, Rustgi SD, Aronson A, Kessel E, Amin S, Lucas AL. Opioid Prescription Is Associated With Increased Survival in Older Adult Patients With Pancreatic Cancer in the United States: A Propensity Score Analysis. JCO Oncol Pract 2022;18:e659–e668.

